# Histopathological evaluation of abdominal aortic aneurysms with deep learning

**DOI:** 10.1101/2024.04.23.24306178

**Authors:** Fiona R. Kolbinger, Omar S. M. El Nahhas, Maja Carina Nackenhorst, Christine Brostjan, Wolf Eilenberg, Albert Busch, Jakob Nikolas Kather

**Affiliations:** Department of Visceral, Thoracic and Vascular Surgery, University Hospital and Faculty of Medicine Carl Gustav Carus, TUD Dresden University of Technology, Dresden, Germany; Else Kröner Fresenius Center for Digital Health (EKFZ), TUD Dresden University of Technology, Dresden, Germany; Weldon School of Biomedical Engineering, Purdue University, West Lafayette, IN, USA; Regenstrief Center for Healthcare Engineering (RCHE), Purdue University, West Lafayette, IN, USA; Department of Biostatistics and Health Data Science, Richard M. Fairbanks School of Public Health, Indiana University, Indianapolis, IN, USA; Department of Pathology, Medical University of Vienna, Vienna, Austria; Division of Vascular Surgery, Department of General Surgery, Medical University of Vienna and Vienna General Hospital, Vienna, Austria; Department of Medicine I, University Hospital Dresden, Dresden, Germany; Department of Medicine III, University Hospital RWTH Aachen, Aachen, Germany; Medical Oncology, National Center for Tumor Diseases (NCT), University Hospital Heidelberg, Heidelberg, Germany

**Author notes:** FRK and OSMEN contributed equally to this work as co-first authors. AB and JNK contributed equally to this work as co-senior authors. Corresponding authors: Albert Busch, Department of Visceral, Thoracic and Vascular Surgery, University Hospital and Faculty of Medicine Carl Gustav Carus, TUD Dresden University of Technology, Fetscherstraße 74, 01307 Dresden, Germany, Jakob Nikolas Kather, Else Kröner Fresenius Center for Digital Health (EKFZ), TUD Dresden University of Technology, Fetscherstraße 74, 01307 Dresden, Germany.

**Keywords:** Abdominal aortic aneurysm, computational pathology, vascular pathology, deep learning

## Abstract

Computational analysis of histopathological specimens holds promise in identifying biomarkers, elucidating disease mechanisms, and streamlining clinical diagnosis. However, the application of deep learning techniques in vascular pathology remains underexplored. Here, we present a comprehensive evaluation of deep learning-based approaches to analyze digital whole-slide images of abdominal aortic aneurysm samples from 369 patients from three European centers. Deep learning demonstrated robust performance in predicting inflammatory characteristics, particularly in the adventitia, as well as fibrosis grade and remaining elastic fibers in the tunica media. Overall, this study represents the first comprehensive evaluation of computational pathology in vascular disease and has the potential to contribute to improved understanding of abdominal aortic aneurysm pathophysiology and personalization of treatment strategies, particularly when integrated with radiological phenotypes and clinical outcomes.

## Introduction

Computational analysis of histopathological imaging has the potential to identify biomarkers, provide insights into disease patterns and pathophysiological processes, and automate histopathological assessment in clinical routine care. The potential of deep learning (DL)-based approaches in vascular pathology has not been characterized in large patient cohorts to date. We evaluated the performance of DL to identify a range of pathological characteristics including inflammation, fibrosis, elastic fiber degradation, angiogenesis, and calcification from digital whole-slide images (WSI) in an expert-annotated dataset of abdominal aortic aneurysm (AAA) wall samples from 369 patients treated at three European centers.

## Methods

### Patient Cohort

Data from a total of 369 patients (84.6% male, mean age 69.1 ± 8.0 years, average maximum diameter 62.9 ± 15.7 mm) undergoing open AAA repair at the Technical University Munich (TUM, n = 287), the University Hospital Würzburg (UHW, n = 36) and the Medical University Vienna (MUV, n = 46), between 2005 and 2019, were included in this study (**Figure 1A**).

**Figure 1:**
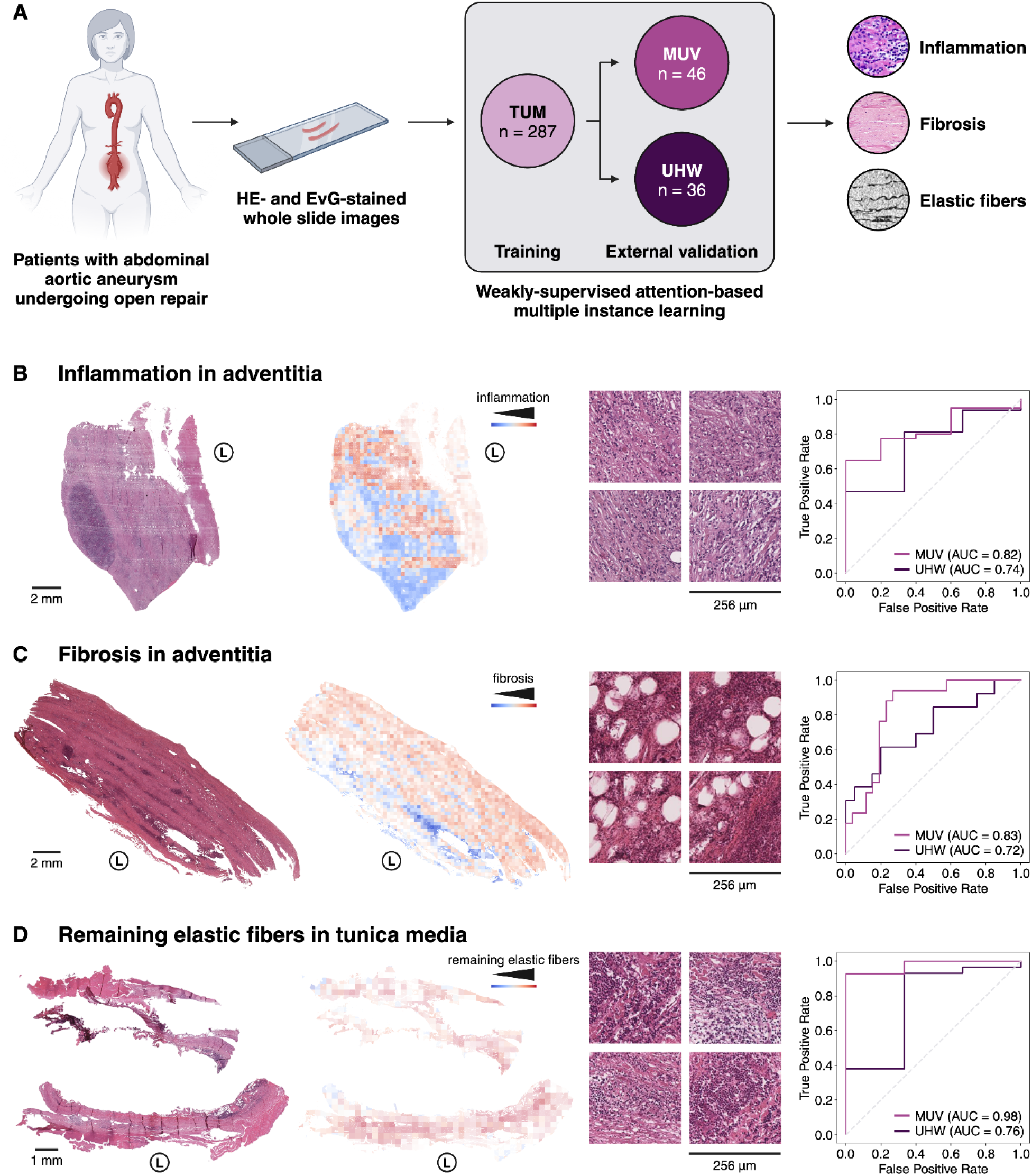
Histopathological evaluation of abdominal aortic aneurysms with deep learning. **(A)** Hematoxylin and Eosin (HE)-stained whole slide images from 369 patients undergoing open aortic aneurysm repair at three European centers were used to develop weakly-supervised deep learning models classifying pathological variables including inflammation **(B)** and fibrosis in the adventitia **(C)**, as well as remaining elastic fibers in the tunica media **(D)**. Model performance was qualitatively evaluated with heatmaps indicating predicted model scores for each 256 μm x 256 μm patch of the whole slide image, as well as with the most predictive patches from the respective slide. Quantitatively, model performance was evaluated using the area under the receiver-operating characteristic curve (AUC) **(B, C, D)**. Adventitial inflammation **(B)** was classified on a four-category ordinal scale (none, minor, intermediate, major inflammation); the figure displays results from a model differentiating none from the other three classes. Adventitial fibrosis **(C)** was classified on a three-category ordinal scale (minor, intermediate, major fibrosis); the figure displays results from a model differentiating major fibrosis from the other two classes. Remaining elastic fibers in the tunica media **(B)** was classified on a binary scale (< 25%, > 25%); the figure displays results from a model differentiating both classes. Abbreviations: Area under the receiver-operating characteristic curve (AUC), Elastica van Gieson (EvG), hematoxylin and eosin (HE), lumen (L), Medical University Vienna (MUV), Technical University Munich (TUM), University Hospital Würzburg (UHW).

Aneurysm samples from the left anterior wall were independently analyzed by three pathologists as described previously^1^ to create ground truth data for model training and testing. In brief, the following histopathological parameters were evaluated: Grade of inflammation in tunica media [none, minor, intermediate, major], grade of inflammation in adventitia [none, minor, intermediate, major], type of inflammation in adventitia and tunica media [none, acute, chronic], Histological Inflammation Scale of Aneurysms (HISA) grade^2^ [0, 1, 2, 3, 4], angiogenesis in tunica media [present, not present], calcification in tunica media [present, not present], grade of fibrosis in adventitia [minor, intermediate, major], remaining elastic fibers in tunica media [< 25%, > 25%]. In case of disagreement, consensus was reached through discussion. Hematoxylin and Eosin (HE)- and Elastica van Gieson (EvG)-stained slides were digitized using an Aperio AT2 (Leica, Wetzlar, Germany) slide scanner. Patient sex and smoking history were gathered as binary clinical parameters.

### Computational models

Weakly-supervised attention-based multiple instance learning models were trained to identify pathological characteristics from HE- and EvG-stained WSI. All models were trained and internally validated by 5-fold cross validation on the TUM cohort and separately tested on external cohorts from MUV and UHW. The STAMP protocol was utilized to process the WSIs^3^. In brief, WSI were preprocessed by tessellation into 224 × 224 pixel patches at a magnification of 256 μm per pixel, followed by computational background rejection, and color normalization of HE-stained slides. EvG-stained slides were not color-normalized as no color normalization protocols exist for this staining. Subsequently, features were extracted using a pre-trained histology image encoder. To aggregate patches for slide-level predictions, downstream classification tasks were modeled through weakly-supervised attention-based multiple instance learning^4^, enabling the correlation of slide-level labels with tissue morphology (**Figure 1A**). Model performance was quantified using the mean area under the receiver operating characteristic (AUC) and standard deviation across 5 folds. High-resolution attention heatmaps for qualitative evaluation were generated through a convolutional equivalent of the attMIL model^4^.

## Results

Based on a multicentric expert-labeled histopathological dataset of resected AAA from 369 patients, we trained DL models to identify histopathological parameters directly from WSI. With regard to inflammatory characteristics, DL could reliably predict presence (binary classification, AUC_MUV_ = 0.75 ± 0.10, AUC_UHW_ = 0.85 ± 0.08) and grade of inflammation (multi-class classification, AUC_MUV_ = 0.73 ± 0.10, AUC_UHW_ = 0.78 ± 0.10) in the adventitia from HE-stained slides (**Figure 1B**). While the models did not include any regional information, areas that were most relevant for prediction of inflammation in the adventitia were indeed localized in the adventitial region of specimens and displayed inflammatory cell infiltration from various cell types (i.e. plasma cells, lymphocytes, **Figure 1B**). In contrast, prediction models for presence and grade of inflammation in the tunica media as well as inflammation type in the adventitia and tunica media yielded random prediction results (AUC overlapping 0.50).

With regard to other pathological characteristics, DL could predict the grade of fibrosis in the adventitia (major vs. other, AUC_MUV_ = 0.83 ± 0.02, AUC_UHW_ = 0.72 ± 0.08, **Figure 1C**) and remaining elastic fibers in the tunica media (AUC_MUV_ = 0.97 ± 0.02, AUC_UHW_ = 0.75 ± 0.12, **Figure 1D**). Furthermore, DL could differentiate HISA^2^ classes (AUC_MUV_ = 0.79 ± 0.10, AUC_UHW_ = 0.77 ± 0.06), historically classified based on the extent of mild and chronic inflammation, lymphocytic infiltration, and fibrosis. Angiogenesis, calcification, sex, and smoking history could not be predicted from HE-stained WSI (AUC overlapping 0.50).

Models trained on EvG-stained WSI overall performed similar to models trained on HE-stained WSI for detection of calcification, fibrosis, HISA grade, sex, and smoking history. For prediction of the abovementioned inflammatory parameters, HE-trained models performed considerably superior to EvG-trained models.

## Discussion

This study presents the largest evaluation of computational pathology in the context of vascular disease to date. Our results provide evidence that DL can reliably predict histopathological features like inflammation, elastic fiber degradation, and fibrosis from HE-stained AAA pathology slides. In addition, DL can predict the HISA grade from unlabeled pathology, affirming the relevance of this pathological classification established in 1994^2^ and the reproducibility of inflammatory characteristics in histologic examination of AAA.

Prior works in computational vascular histopathology have evaluated basic segmentation tasks in smaller, monocentric patient cohorts^5,6^. Previous DL-based studies in AAA have predominantly focused on aneurysm detection and segmentation or prediction of clinical endpoints such as aneurysm growth or rupture risk from radiological or biomechanical data.^7,8^ Given the nature of AAA and the impracticability of aortic sampling, our findings cannot provide directly actionable clinical insights. However, our results have the potential to contribute to improved understanding of AAA biology and treatment, especially when set in relation to other data modalities (i.e., radiological imaging, clinical risk factors) in multimodal prediction models.

Our finding of computationally discernible inflammatory patterns in the adventitia implies potential for non-invasive diagnostic approaches: Imaging techniques like PET, SPECT, and radiolabeled white blood cell scintigraphy can non-invasively assess inflammation levels in the adventitia^9^ and could potentially identify patients who could benefit from pharmacotherapy targeting inflammation^10^. To maximize translational impact, our work warrants linking of AAA histopathological phenotypes with respective radiological and biomechanical phenotypes as well as clinical outcomes.

## Declarations

### Abbreviations

AAA: Abdominal Aortic Aneurysm
AUC: Area under the receiver operating characteristic
EvG: Elastica van Gieson
HE: Hematoxylin and Eosin
HISA: Histological Inflammation Scale of Aneurysms
MUV: Medical University Vienna
TUM: Technical University Munich
UHW: University Hospital Würzburg
WSI: Whole-slide image

## Data Availability Statement

Due to data privacy regulations, the raw imaging data cannot be published along with this work. The feature vectors extracted from the histopathological WSI and the corresponding clinical endpoints analyzed in this work are available at https://doi.org/10.5281/zenodo.10998463. Code for preprocessing is available at https://github.com/KatherLab/STAMP. Code for modeling is available at https://github.com/KatherLab/marugoto. Code for spatial heatmaps and top-attention tiles is available at https://github.com/KatherLab/highres-WSI-heatmaps/tree/AAA_heatmaps.

## Author Contributions

FRK: Conceptualization, Data curation, Formal Analysis, Investigation, Methodology, Validation, Visualization, Writing – original draft, Writing – review & editing; OSMEN: Conceptualization, Data curation, Formal Analysis, Investigation, Methodology, Software, Validation, Visualization, Writing – original draft, Writing – review & editing; MCN: Data curation, Formal Analysis, Validation, Visualization, Writing – review & editing; CB: Data curation, Formal Analysis, Validation, Visualization, Writing – review & editing; WE: Data curation, Formal Analysis, Validation, Writing – review & editing; AB: Conceptualization, Data curation, Formal Analysis, Funding acquisition, Project administration, Resources, Supervision, Validation, Visualization, Writing – review & editing; JNK: Conceptualization, Funding acquisition, Methodology, Project administration, Resources, Supervision, Writing – review & editing.

## Ethical Approval

This study was performed in accordance with the declaration of Helsinki and its later amendments. The local institutional review boards reviewed and approved the original data collection (Technical University Munich: #2799/10 and #576/18S, University Hospital Würzburg: #188/11, Medical University Vienna: #1729/2014). The overall analysis was approved by the ethics committee of the Technical University Dresden (BO-EK-444102022). All patients had a clinical indication for open AAA repair and gave informed consent to data collection and analysis.

## References

1. Busch, A. Abdominal aortic aneurysms harbor different histomorphology not associated with classic risk factors, the HistAAA study. medRxiv 2024.04.16.24305904 (2024) doi:10.1101/2024.04.16.24305904.

2. Rijbroek, A., Moll, F. L., von Dijk, H. A., Meijer, R. & Jansen, J. W. Inflammation of the abdominal aortic aneurysm wall. Eur. J. Vasc. Surg. 8, 41–46 (1994).

3. El Nahhas, O. S. M. et al. From Whole-slide Image to Biomarker Prediction: A Protocol for End-to-End Deep Learning in Computational Pathology. arXiv [cs.CV] (2023).

4. El Nahhas, O. S. M. et al. Regression-based Deep-Learning predicts molecular biomarkers from pathology slides. Nat. Commun. 15, 1–13 (2024).

5. Thorsted, B. et al. Artificial intelligence assisted compositional analyses of human abdominal aortic aneurysms ex vivo. Front. Physiol. 13, 840965 (2022).

6. Niemann, A., Talagini, A., Kandapagari, P., Preim, B. & Saalfeld, S. Tissue segmentation in histologic images of intracranial aneurysm wall. Interdisciplinary Neurosurgery 26, 101307 (2021).

7. Lu, J.-T. et al. DeepAAA: clinically applicable and generalizable detection of abdominal aortic aneurysm using deep learning. arXiv [eess.IV] (2019).

8. Kim, S. et al. Deep Learning on Multiphysical Features and Hemodynamic Modeling for Abdominal Aortic Aneurysm Growth Prediction. IEEE Trans. Med. Imaging 42, 196–208 (2023).

9. Husmann, L. et al. Imaging characteristics and diagnostic accuracy of FDG-PET/CT, contrast enhanced CT and combined imaging in patients with suspected mycotic or inflammatory abdominal aortic aneurysms. PLoS One 17, e0272772 (2022).

10. Miyake, T. & Morishita, R. Pharmacological treatment of abdominal aortic aneurysm. Cardiovasc. Res. 83, 436–443 (2009).

